# COVID-19 Transmission Within Danish Households: A Nationwide Study from Lockdown to Reopening

**DOI:** 10.1101/2020.09.09.20191239

**Authors:** Frederik Plesner Lyngse, Carsten Thure Kirkeby, Tariq Halasa, Viggo Andreasen, Robert Leo Skov, Frederik Trier Møller, Tyra Grove Krause, Kåre Mølbak

## Abstract

**Background:** The Covid-19 pandemic is one of the most serious global public health threats in recent times. Understanding transmission of SARS-CoV-2 is of utmost importance to be able to respond to outbreaks and take action against spread of the disease. Transmission within the household is a concern, especially because infection control is difficult to apply within the household domain.

**Methods:** We used comprehensive administrative register data from Denmark, comprising the full population and all COVID-19 tests, to estimate household transmission risk and attack rate.

**Results:** We studied the testing dynamics for COVID-19 and found that the day after receiving a positive test result within the household, 35% of potential secondary cases were tested and 13% of these were positive. After a primary case in 6,782 households, 82% of potential secondary cases were tested within 14 days, of which 17% tested positive as secondary cases, implying an attack rate of 17%. Among primary cases, those aged 0-24 were underrepresented when compared with the total population. We found an approximately linearly increasing relationship between attack rate and age. We investigated the transmission risk from primary cases by age, and found an increasing risk with age of primary cases for adults, while the risk seems to decrease with age for children.

**Conclusions:** Although there is an increasing attack rate and transmission risk of SARS-CoV-2 with age, children are also able to transmit SARS-CoV-2 within the household.

## 1 INTRODUCTION

In late 2019, increased numbers of severe respiratory infections were reported in the Wuhan region in China caused by the Sars-CoV-2 virus (Coronaviridae Study Group of the International Committee on Taxonomy of Viruses, 2020).

Since its emergence, the virus has spread rapidly throughout all five continents affecting millions of people (World Health Organization, 2020) and causing massive economic losses (Fernandes, 2020). The effective reproduction number was estimated in different studies with different methods to range from 2.1 to 4.7 (Boldog et al. (2020)), revealing a high transmission potential. Person-to-person transmission is a major transmission mode of SARS-CoV-2, including transmission through aerosols or droplets on surfaces (Chan et al., 2020; Leclerc et al., 2020). Quantifying the transmission risk in different settings is essential for improving our understanding of the viral transmission dynamics, to implement effective preventive measures, to minimize economic damage, and to avoid overloading the healthcare system. Close person-to-person contact is one of the main risk factors and transmission within the household is thus a major setting for virus transmission (Prem et al., 2017; Davies et al., 2020; Cauchemez et al., 2009; Leclerc et al., 2020). Furthermore, infection control and isolation are challenging in the (often crowded) household domain. Quantifying the extent of transmission within the household can help improve our understanding of the effects of implementing quarantine for household members, physical distancing and improved hygiene. Furthermore, these estimates are useful in constructing reliable prediction models for the spread of SARS-CoV-2 from households to the community.

Data from contact tracing and monitoring of individuals have been used to investigate household transmission of SARS-CoV-2 (He et al., 2020; Jing et al., 2020; Liu et al., 2020; Li et al., 2020; Madewell et al., 2020). Contact tracing is laborious and requires large resources when there are many new cases, as in the case of the SARS-CoV-2 pandemic. Thus, these studies have been limited to include a maximum of a few hundred primary cases, selected mainly due to history of hospitalization or clinical disease (see a systematic review by Madewell et al. (2020)). The relatively small sample size as well as the selection and recall bias caused by contact tracing may limit the generalizability of these studies. In this study, we exploit national register data from Denmark to investigate transmission patterns within households. To our knowledge, this is the first nationwide study that uses estimates of household attack rates and transmission risks that exploit SARS-CoV-2 test data from an entire population.

## 2 DATA AND METHODS

### 2.1 Data and summary statistics

#### The Epidemic in Denmark

On February 27, 2020, the first case of SARS-CoV-2 was diagnosed in Denmark (Reilev et al., 2020). Shortly thereafter, the number of cases began to rise with an estimated effective reproduction rate (*Re*) of about 2.5 (Statens Serum Institut (2020)). On March 11, a comprehensive lockdown of the public sector was implemented by the government. Moreover, the private sector was encouraged to work from home as much as possible. In Figure A.1 (panel a), key indicators of the epidemic are shown: number of tests, positive test results, hospitalized cases, and deaths for the early stage of the epidemic, the lockdown period and the two following stages of reopening. The figure shows a clear reduction in the number of positive test results, hospitalized cases and deaths over time. Panel b shows a summary of the main measures for controlling the epidemic over time, including test and contact tracing strategy, and restrictions on public and private workplaces, education and childcare institutions, as well as in the community in general.

All Danish residents have access to tax-paid universal health insurance, and a test for SARS-CoV-2 is free of charge. Furthermore, Denmark has comprehensive social welfare insurance, and SARS-CoV-2 sick leave is fully reimbursed by the state. Thus, financial reasons are not a major obstacle to obtaining a test. The test capacity has increased throughout the epidemic, and in the study period, the number of tests has been fairly stable since late April 2020 (Figure A.1). In the beginning of the epidemic, all suspected cases of COVID-19 were tested and their contacts traced. However, soon after, due to capacity constraints on testing, only cases with severe symptoms (i.e., admitted to hospital) were tested and the overstretched contact tracing was halted. During reopening testing again became generally accessible, so that all residents could obtain a test without a referral (Panel b of Figure A.1).

**Figure 1.**
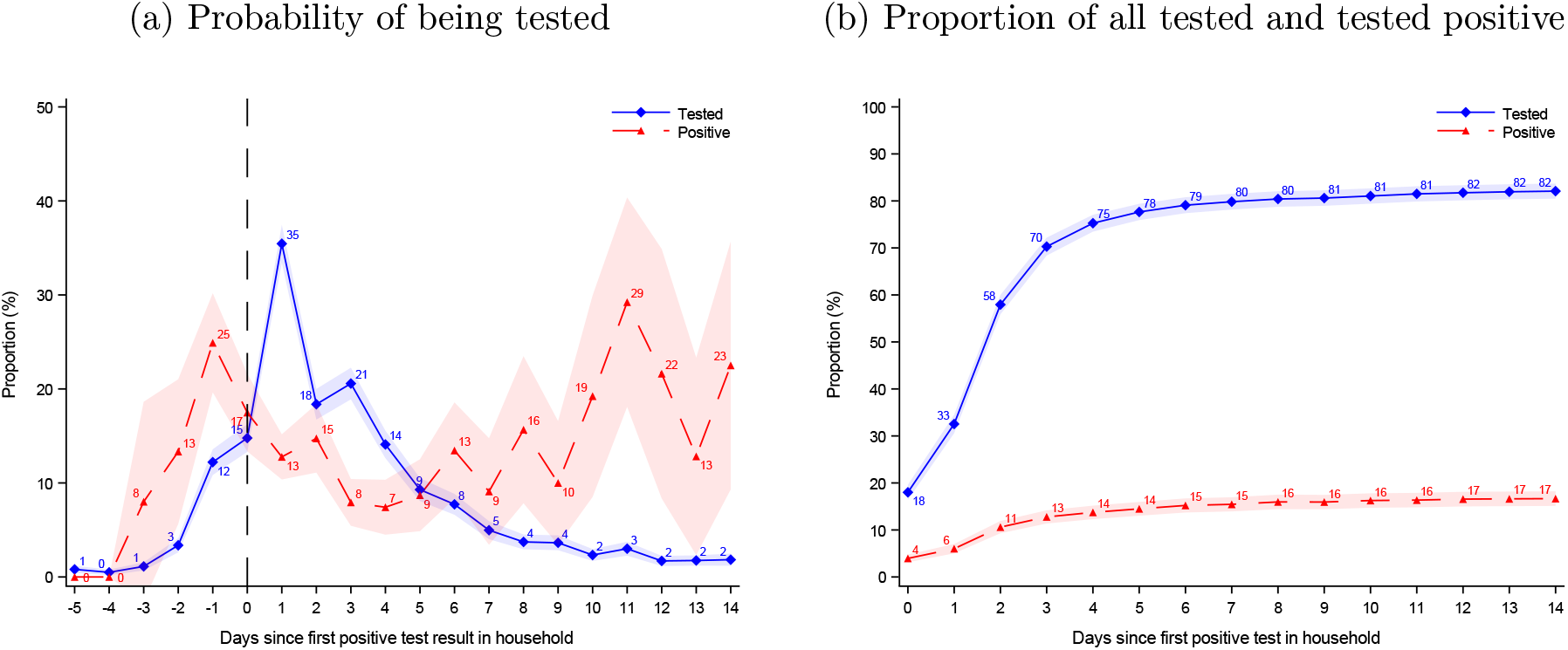
Testing dynamics

Notes: Data shows period 3: Late Reopening. Results for the three periods are presented in Figure D.1. In Panel a, *t* = 0 denotes the time of receiving the first positive test result in a household. On the day preceding the result (*t* = *−*1) 12% were tested and 25% of these tested positive. The day after the households received their first positive test result (*t* = 1), 35% were tested and 13% of these tested positive. In Panel b, *t* = 0 denotes the time the first positive test was taken (not when the test result was given). 18% of all potential secondary cases were tested on the same day as the primary case (*t* = 0) and 4% of all potential secondary cases tested positive. This implies a 22% probability of testing positive conditional on being tested on the same day as the primary case. 14 days after the day of the first positive test, 82% had been tested and 17% tested positive. Shaded areas show the 95% confidence bands clustered on household level.

#### Register data

In the current study, we used Danish administrative register data. All residents in Denmark have a unique personal identification number that allows a completely accurate linkage of information across different registers at the individual level. All microbiological data in Denmark is registered in the Danish Microbiology Database, from where we obtained individual level data on all national tests for SARS-CoV-2 for the period February 27, 2020 (the first positive test in Denmark) to July 24, 2020. Information on the reason for being tested (e.g., symptoms, potential contact with infected persons etc.) was not available. From the Danish Civil Registry System, we obtained information about the sex, age, and home address for all individuals living in Denmark. All data management and analyses were carried out on the Danish Health Data Authority’s restricted research servers with project number FSEID-00004942.

#### Data linkage

We constructed households by linking all individuals living at the same address, and only considered households with six or fewer members, in order to exclude e.g., institutions. The data set captured 98.3% percent of the Danish population. Person level data, which included information on the test result and date and time of sampling as well as time of the result, were linked to individuals within households. For each household, we identified the first positive test for SARS-CoV-2; this case was defined as the primary case and referred to as such throughout this paper. We considered all subsequent tests from other members in the same household as tests taken in response to the primary case. We defined secondary cases as those who had a positive test within 14 days of the primary case being tested positive. Primary examination of the data revealed that this cutoff provided a stable proportion of potential secondary cases (see Figure 1b). In addition, we assumed that the secondary household members were infected by the household primary case, although some of these secondary cases could represent co-primary cases. A longer cutoff time period could result in misclassification of cases among household members with somewhere else being the source of secondary infections.

#### Robustness of estimates over time

In order to investigate potential bias in our results in relation to time, due to changing test strategy and capacity over the epidemic period (see Figure A.1 for an overview of test strategy and capacity), we separated the data into three data sets representing the three time periods based on the test date of the primary case, and analyses were performed separately on these data sets. The defined periods were: Lockdown: March 12 – April 14; Early reopening: April 15 – May 17; and Late reopening May 18 – July 5 (see Figure A.1). During lockdown, educational institutions were kept closed and the public sector with non-essential functions stayed at home. Children’s daycare was limited to children of employees in essential functions, such as doctors, nurses and police. Employees in the private sector were encouraged to stay home if possible, and international travel was minimized by closing the borders except for essential activities. In the early reopening phase, children’s daycare became available again, and schools were re-opened for classes up to 5th grade. In the late reopening, systematic contact tracing was resumed, schools (6-10th grade) and higher educational institutions reopened, along with restaurants and smaller bars (with physical distancing).

### 2.2 Methods

To determine the probability that an additional household member would test positive after the primary case in the household was tested positive, we used all records for potential secondary cases within the household.

We defined the *attack rate* as the proportion of additional household members that tested positive, whereas the *transmission risk* was the proportion of secondary cases per primary case. We did not exclude any potential co-primary cases in the main analysis. We further conducted a sensitivity analysis for the robustness of the time cutoff of transmission from the primary case to potential secondary cases.

We used SAS 9.4 to manage and analyze the data (SAS, 2015). In order to reach sufficient sample sizes, we separated all records into age groups of five-year intervals.

#### 2.2.1 Testing Dynamics

To investigate the testing dynamics, we took an event study approach (Ball & Brown, 1968). Following this method, we used the date of diagnosis of the primary case in each household as an event and observed all other household members for five days before until 14 days after the event. In the case of two or more primary cases detected on the same date, we randomly assigned one of them as the primary case. We estimated the probability of being tested (*β_τ_*) for each day relative to the first positive test result within the household, using the following equation:

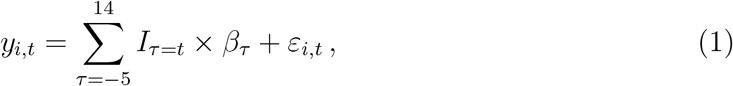

where *y_i,t_* is a binary variable for individual *i* being tested at time *t. τ* is days relative to the date of the primary case’s positive test result. *I_τ=t_* denotes indicators for time since the primary case’s positive test result. *β_τ_* represents parameters estimating the probability of being tested on day *τ* relative to receiving the primary case’s test result in the household. *ε_i,t_* denotes the error term, clustered on the household (event) level. We used the same equation to estimate the probability of *y* testing positive conditional on being tested.

Furthermore, to estimate the proportion of potential secondary cases that had been tested on day *τ* or previously, we estimated the absorbing probability, using the following equation:

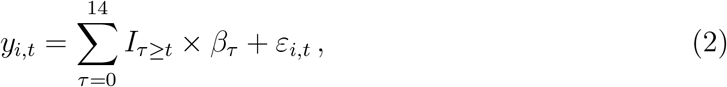

where *I_τ≥t_* = 1 if individual *i* was tested on day *τ* or previously, and zero otherwise. We also used the same equation to estimate the probability of *y* ever being tested positive.

#### 2.2.2 Proportion of Total Positive Cases Originating from Households

To investigate the proportion on positive tests originating from households, we defined new cases that live in a household with another case that tested positive within the preceding 14 days as a case originated from the household domain. We used a seven day rolling average in order to take account of variation in testing rates across the weekdays.

#### 2.2.3 Attack rate

To estimate the attack rate, we estimated the proportion of potential secondary household members who received a positive test within 14 days after the test date of the primary case. We estimated attack rates using the following equation:

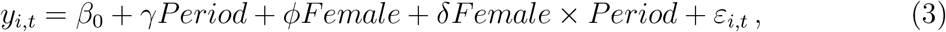

where *y_i,t_* = 1 if the individual had a positive test within the 14 days after the primary case, and zero otherwise. *γ* denotes a vector of fixed effects for the three periods. Female is a binary variable for sex. *β*_0_ measures the 14 day attack rate. *ε_i,t_* denotes the error term, clustered on the household (event) level.

#### 2.2.4 Age Structured Attack Rate and Transmission Risk

To investigate the age structure of the attack rate for potential secondary cases and transmission risk from primary cases, we used a non-parametric approach. We separated the data into five-year age groups. We estimated the attack rate using the following equation:

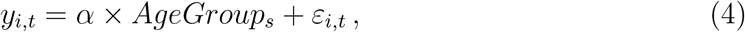

where *y_i,t_* = 1 if the potential secondary case *i* had a positive test within the 14 days after the test date of the primary case, and zero otherwise. *AgeGroup_s_* is five-year age groups of the potential secondary cases. *α* is a vector that measures the age structured attack rate. *ε_i,t_* denotes the error term, clustered on the household (event) level.

We estimated the transmission risk using the following equation:

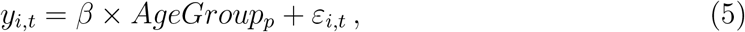

where *AgeGroup_p_* is five-year age groups of the primary cases. *β* is a vector that measures the age structured transmission risk.

We estimated the age structured interaction between the attack rate and transmission risk using the following equation:

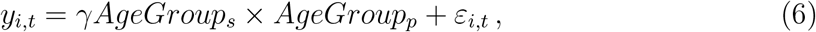

where *γ* is a vector that measures the age structured interaction between attack rate and transmission risk.

To quantify the effect of age on attack rate, we estimated the approximately linear relationships between attack rate and age using equation:

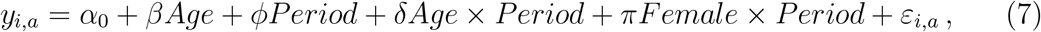

where *β* measures the probability of being infected as a linear function of age. *φ* is a vector of fixed effects for each time period (see “Robustness of estimates over time” in section 2.1). *δ* measures the differential age gradient for each period. *π* measures the effect of sex for each period.

In order to investigate the potential difference between male and female cases, we explored the age dependent attack rate separately for each sex. We also separated the data into households where the primary cases were children (below 15 years of age) and adults (above 25 years of age). These thresholds were chosen to ensure the primary cases were either children or adults.

#### 2.2.5 Household Size Structured Attack Rate

We estimated the attack rate stratified by the number of household members for households with one to six members. Furthermore, we estimated the proportion of households with N number of positive cases, conditional on the size of the household being greater than or equal to N (with a maximum household size of six).

## 3 Results

### 3.1 Descriptive statistics

In total, we obtained positive test results from 6,782 household primary cases and 1,904 positive secondary cases, see Table 1. Further summary statistics are presented in Appendix B.

**Table 1:**
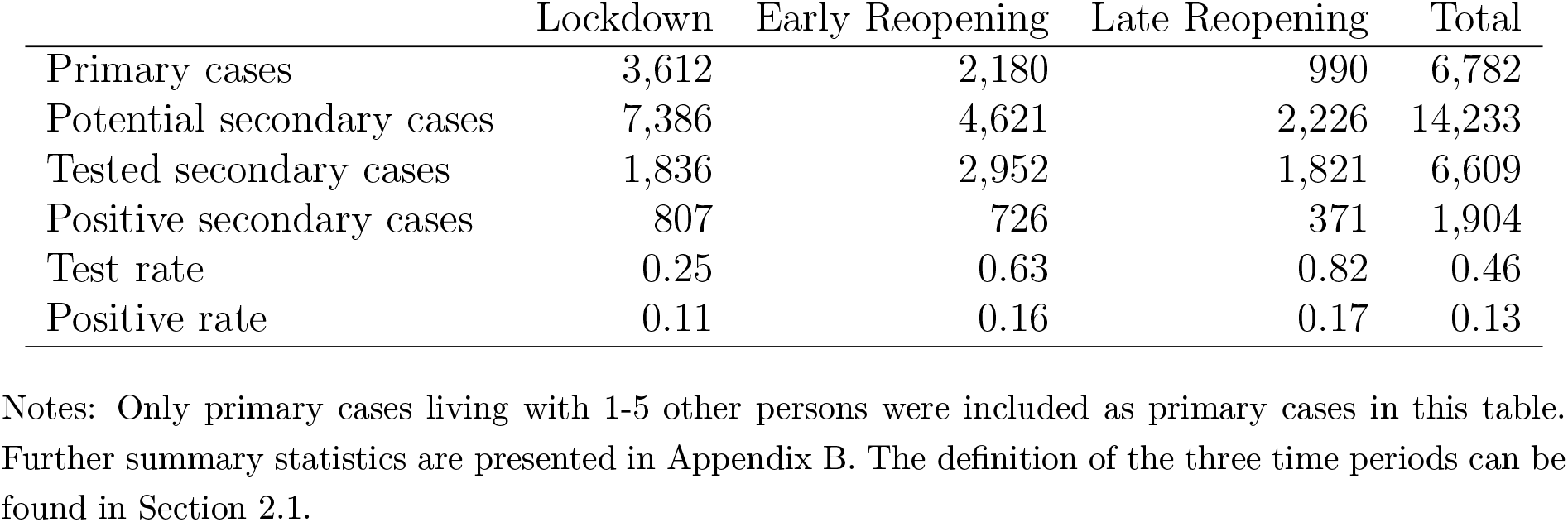
Summary Statistics of the Study Population

|  | Lockdown | Early Reopening | Late Reopening | Total |
| --- | --- | --- | --- | --- |
| Primary cases | 3,612 | 2,180 | 990 | 6,782 |
| Potential secondary cases | 7,386 | 4,621 | 2,226 | 14,233 |
| Tested secondary cases | 1,836 | 2,952 | 1,821 | 6,609 |
| Positive secondary cases | 807 | 726 | 371 | 1,904 |
| Test rate | 0.25 | 0.63 | 0.82 | 0.46 |
| Positive rate | 0.11 | 0.16 | 0.17 | 0.13 |

Notes: Only primary cases living with 1-5 other persons were included as primary cases in this table. Further summary statistics are presented in Appendix B. The definition of the three time periods can be found in Section 2.1.

Appendix figure A.2 presents the probability of being tested and testing positive over the epidemic for five-year age groups. We found that all age groups are being tested, though children generally had a lower probability. The increase in test capacity over time was approximately equally distributed across ages. In late June, we saw an increase in the probability of children being tested.

Appendix figure B.1 shows summary statistics of the primary cases compared to the overall Danish population. Panel (a) shows the age distribution and Panel (b) shows the household size distribution. There were proportionally fewer children as primary cases than in the total population. The household sizes of the primary cases generally matched the household sizes of the population.

### 3.2 Testing Dynamics

In this section, we focus only on the testing dynamics during the late reopening, where test capacity was stable. (In Appendix D, we illustrate changes over all three periods.)

Figure 1 panel (a) shows that after receiving a positive test result in the household (*t* = 0), 36% of potential secondary cases were tested (blue) the day after the positive test result (*t* = 1) of the primary case was available and 13% of these 36% were positive (red). On the day preceding the test result (*t* = *−*1), 12% were tested and 24% of these tests were positive.

Panel (b) shows the proportion of individuals that were tested (blue) and those that tested positive (red), daily up to 14 days after the primary case was tested (*t* = 0). 18% of potential secondary cases were tested on the same day as the primary case and 4% of potential secondary cases tested positive on that day. Within 14 days after the primary case was tested, 82% of the potential secondary cases were tested and 17% tested positive.

There were 4% who tested positive on the same day (*t* = 0) as the primary case. These may represent co-primary cases, and therefore do not represent household cases. Under this assumption, in order to estimate the attack rate, one should exclude these cases (by subtracting 4 percentage points (pp) from 17%), thereby resulting in an attack rate of 13%. Similarly, cases detected within one day of the primary case may represent co-primary cases (*t ≤* 1). This leaves an attack rate of 11% (by subtracting 6 pp from 17%). (Appendix F provides robustness analysis on the definition of co-primary cases.)

#### Proportion of Total Positive Cases Originating from Households

Figure 2 shows the proportion of positive tests originating from households within the 14 days after the primary case tested positive. After a rapid decrease immediately after lockdown, there was an increasing proportion of new cases originating from households during lockdown, and a decreasing proportion after reopening. The last day of school was June 26, and many families start their vacation at that time. After this date, the proportion started to increase again.

**Figure 2:**
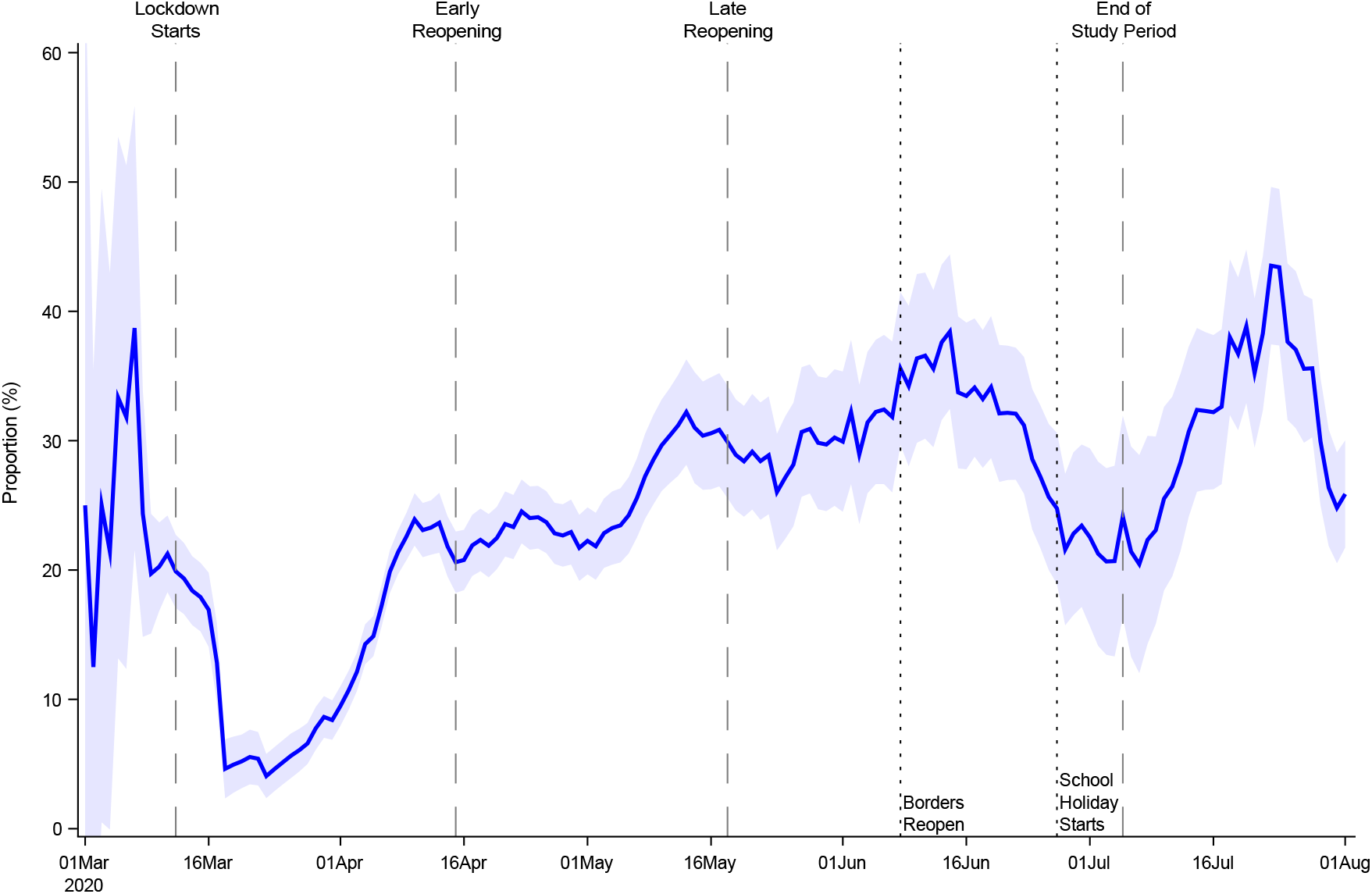
Proportion of Total Positive Cases Originating from Households

Notes: This figure shows the proportion of positive tests originating from households, as defined by new cases that live in a household with another case that tested positive within the preceding 14 days. The figure shows a seven day moving average, while the shaded area shows the 95% confidence bands with standard errors clustered on the individual level.

### 3.3 Age structured attack rate

We found an approximately linearly increasing relationship between attack rate and age. Panel a of Figure 3 shows the probability of having a test (blue) and the probability of having a positive test (red) (unconditional on being tested) across 5-year age groups. The shaded areas show the 95% confidence bands. Panel b shows the transmission risk from primary cases by age group, i.e., the probability of infecting others. The figure shows an increase with age for adults, while the risk seems to decrease with age for children.

**Figure 3:**
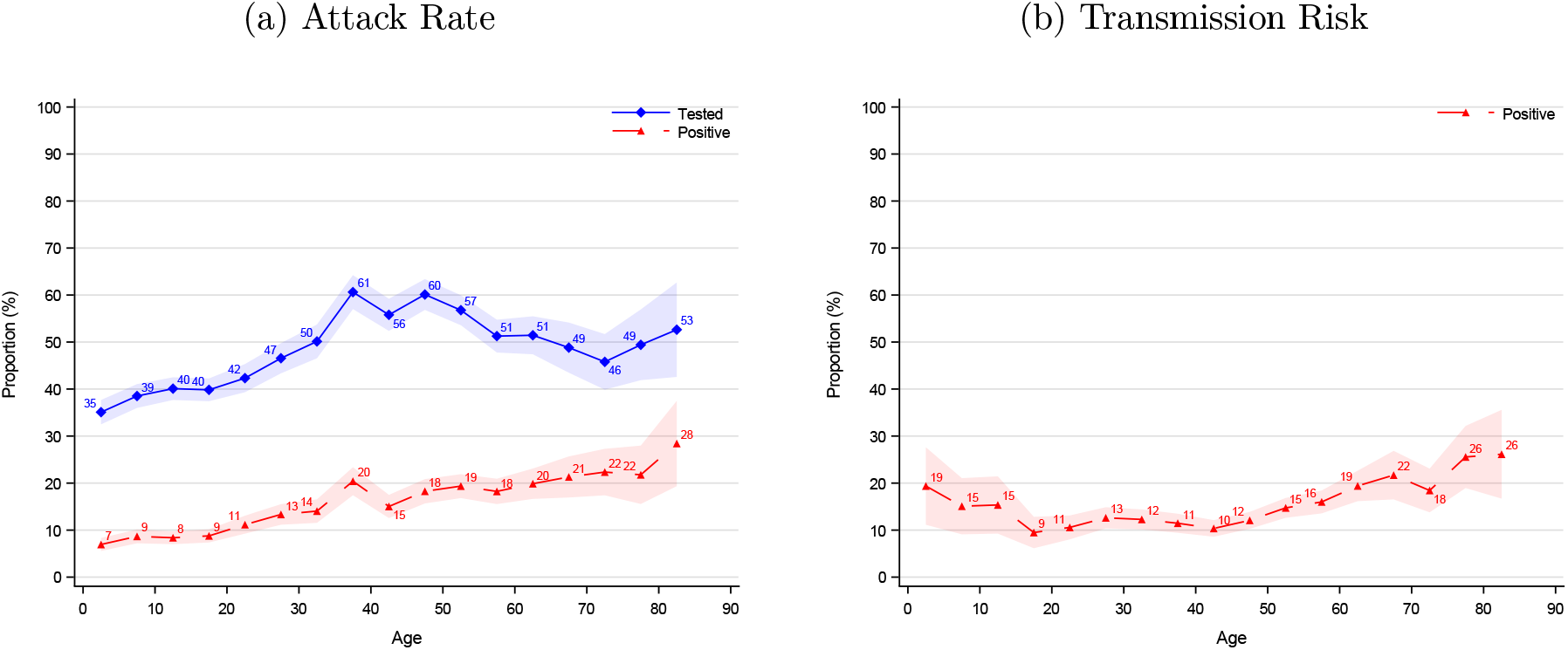
Age Structured Attack Rate and Transmission Risk

Notes: Panel a illustrates the probability of having a test (blue) and the probability of having a positive test (red) (unconditional on being tested) across 5-year age groups. Panel b illustrates the transmission risk from primary cases by age group, i.e. the probability of infecting others. The shaded areas show the 95% confidence bands clustered on the household level.

As figure 3 shows an approximately linear relationship between attack rate and age, we estimated the linear relationship using Equation 7 (see Table E.2, column I). The results show that individuals had a baseline risk of 9.4% of testing positive. The risk increased by 0.24 percentage points for each year of age. Thus, a 10-year-old had a risk of 11.8%, a 30-year-old had 19.0%, and a 60-year-old had 33.4%. The estimates were robust to different specifications, including period and sex covariates, Table E.2, column II-IV.

The attack rates conditional on the primary case being a child or an adult are presented in Figure C.1. When the primary case is a child (under 15 years old), the attack rate seemed uniform, regardless of the age of the potential secondary case. When the primary case is an adult (over 25 years old), the attack rate increased (linearly) with age.

The attack rates by sex are presented in Appendix C.1. The results indicated no difference in attack rates between males and females.

The interaction between the attack rate and transmission risk for each combination of age groups is presented in Figure C.2. The age range of the primary case is depicted on the horizontal axis and the age of potential secondary cases on the vertical axis. Panel a shows the probability of being tested (for each age group combination); Panel b shows the probability of obtaining a positive test result.

The probability for potential secondary cases of being tested was generally highest when the primary case was a child (below 15 years old). The probability of being tested was also high when the age difference between the primary case and the potential secondary case was small. The attack rate was highest when the primary and potential secondary cases were above 60 years of age. When children were the primary case, an increased attack rate was observed for all age groups.

### 3.3.1 Proportion of Cases by Household Size

Figure 4 illustrates the proportion of cases by the number of household members for households with one to six members. For instance, in a household with two members (red), 79% of the households had one positive case, while 21% had two positive cases. Similarly, in a household with three members (green) 79% of the households had one case, 16% had two cases, and 5% had three cases. (Table E.4.) This implies that in a three-person household, 79% have an attack rate of 0, 16% have an attack rate of 50%, and 5% have an attack rate of 100%. This amounts to an overall attack rate of 13% (79% *×* 0% + 16% *×* 50% + 5% *×* 100% = 13%).

**Figure 4:**
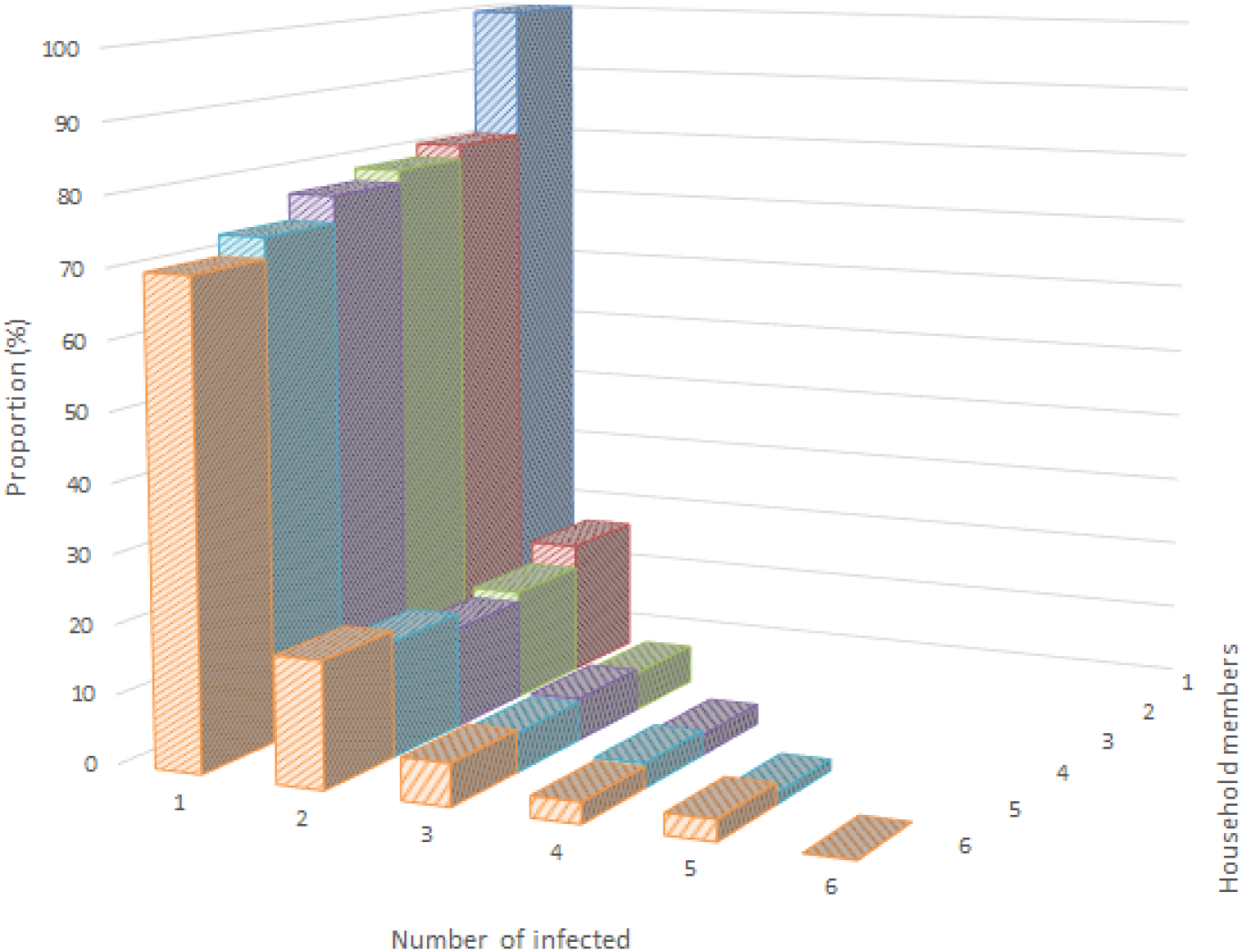
Proportion of Cases by Household Size

Once a primary case was found within a household, the probability of at least one secondary case was 23%, regardless of household size, and consequently 77% of the primary cases do not generate any additional cases. In particular, the primary cases had a 17% probability of generating one additional case, 6% of generating two additional cases, 3% of generating three additional cases, and 2% of generating four additional cases. This pattern was consistent regardless of the number of persons in the household, showing a transmission pattern that was exponentially decreasing with the number members within the household. The pattern was consistent over the three phases of the epidemic examined here, indicating that it is not a result of the change in the testing strategy (Figure D.3).

## 3.4 Robustness of Estimates

The epidemic in Denmark changed over time, both due to changes in policy response (e.g., lockdown), and changes in test capacity and strategy (see Appendix A for an overview). In this section, we investigate the robustness of the previously described results over the three defined periods of the epidemic (see Appendix D). The analyses show that despite the substantial changes in probability of being tested, the estimated attack rates were consistent over the epidemic.

In Appendix F, we investigate the sensitivity of the estimated attacks to the definition of co-primary cases. The analysis showed that the estimated age structured attack rates did not change noticeably by excluding secondary cases found within the same day, one day or two days of the primary case.

## 4 Discussion and Conclusion

We estimated an overall attack rate within households of 17% (Table E.1), ranging from 11% during lockdown, 16% during early reopening, and 17% during late reopening (Figure D.1). This suggests that attack rates estimated early in the Danish epidemic underestimated the true attack rate. This bias likely comes from the limited testing capacity early on. These estimates are in line with the estimates from the literature; Madewell et al. (2020) conducted a meta-analysis of household transmission of SARS-CoV-2 from 40 studies and found an overall attack rate of 18.8%, which is very close to our estimate.

We studied the testing dynamics for COVID-19 and found that the probability of obtaining a test relative to a primary case positive test result within the household peaked on the day after the primary case received the result, where 35% of all potential secondary cases were tested and 13% of these were positive (Figure 1a). Interestingly, 12% of the potential secondary household cases were tested one day preceding the test result of the primary case, and 25% of these were positive. This could indicate that these individuals were tested for a reason other than the primary case result, e.g., for having symptoms themselves. The probability of being tested after a primary case in the same household increased from 18% on the same day as the primary case until it flattened out at around 80% on day six (Figure 1b). 76% of the secondary cases were found during the first three days after the primary case (13% / 17% = 76%). This highlights the importance of fast contact tracing, as most secondary cases are found in the first couple of days after the primary case, which is also concluded by Moghadas et al. (2020).

The proportion of positive cases originating from households (Figure 2) increased during the lockdown until the late reopening period when the borders reopened. Thereafter, it increased again after the school holidays started. In other words, the school holidays (which also include three-weeks of annual leave for most parents) essentially functioned as another lockdown period, because families tend to stay together during the holidays. We suggest that this may have contributed to a low incidence of community transmission over the summer. When the testing capacity was relatively stable (from late April 2020), the proportion varies between 20% and 45% of total cases. This indicates that many cases originate within the household domain, and this should be taken into account when monitoring the epidemic as well as in national guidelines for COVID-19 prevention. Evidence from other countries also finds a substantial proportion of cases originating from households, as for instance, in Israel, where 67% of all infections were found to have been originated at home (Jaffe-Hoffman, 2020).

We estimated the age structured attack rate and transmission (Figure 3) and found a (linear) relationship between age and both the attack rate and transmission risk from primary cases. This suggests that susceptibility to infection increases with the age of the susceptible person. However, children (younger than 15 year of age) also have an elevated transmission risk, likely due to closer contact with parents (Figure C.2), indicating that children may represent an overlooked risk. Our findings correspond well with findings in the literature: Madewell et al. (2020) found that susceptibility to infection increased with age, and a large seroprevalence study in Spain also found an increasing linear relationship with age (Pollán et al., 2020). Similar findings were found by Li et al. (2020) and Bi et al. (2020).

We further estimated the attack rates conditional on the primary case being a child or an adult (Figure C.1). When the primary case is a child (under 15 years old), the attack rate seemed to be uniform, regardless of the age of the potential secondary case. When the primary case is an adult (over 25 years old), the attack rate increased (linearly) with the age of the potential secondary case. This suggests that transmission from children is fairly constant, and depends on close contact with susceptible cases, whereas transmission from adults is more effective the older the potential secondary case is. One could think that if a child is sick, caregivers are likely to have more even close contact with the child – and more so the younger the child is. The opposite may be true for adult cases, indicating that the susceptibility to COVID-19 increases with the age of a person, reflecting immunological properties. Although there is agreement that transmission from and between children is not the main driver in this epidemic (Ludvigsson, 2020), transmission from sick children to parents in the household domain may represent a hitherto overlooked risk factor.

We also estimated the age structured attack rates by sex and found no difference in attack rates between males and females, corresponding to the findings of Jing et al. (2020).

We used a large administrative data set to investigate the attack rate and transmission risk from lockdown to reopening, comprising 6,782 primary cases and 14,233 potential secondary cases. Exploiting this rich nationwide data, we were able to address several concerns regarding the sensitivity of our results.

As the testing capacity and strategy (and hence access to obtaining a test) has changed remarkably over the epidemic, robustness of results are a primary concern when comparing positive test cases over the COVID-19 epidemic. We addressed this by dividing our sample into three periods with different testing capacities (Appendix D) and found that the probability of obtaining a test did increase remarkably across the periods. Our results were, however, relatively consistent over time, suggesting that the findings are not due to changing testing strategies.

There are no formal guidelines for defining co-primary cases. Madewell et al. (2020) provided a review on household transmission of SARS-CoV-2: most of the studies included in Madewell et al. (2020) did not describe how co-primary cases were handled, several studies stated they assumed all secondary cases were infected by the primary case, one study excluded secondary cases if they developed symptoms before exposure to the primary case, and another study randomly selected one primary case as the source of infection. We addressed this important question by showing the sensitivity for different specifications. We found that the attack rate is strongly dependent on the definition. For instance, in Figure 1b we show that if one defines co-primary cases as individuals tested on the same day as the primary case, the secondary attack rate is reduced by 24% (4% / 17% = 24%). Increasing the period for co-primary cases further caused the estimated attack rate to decrease even more. Therefore, it is important to further investigate the dynamics of SARS-CoV-2 in homes, schools, workplaces and major places in the community in order to quantify their impacts on virus transmission and develop effective control measures.

Mathematical modeling is a widely used tool for researchers in order to understand and predict the spread of disease; and policy relies on proper results from these models when making decisions such as choosing between keeping some parts of society open and other parts closed. Closing of childcare and schools has been widespread in most countries. The results from this study can be used as direct input in parameterizing such mathematical models in terms of virus transmission at home. Furthermore, we estimated the age structured attack rate and transmission risk, as well as the interaction of these, which are important inputs in mathematical models, for instance, for contact matrices between age groups (Davies et al., 2020; Zhang et al., 2020).

When modeling the spread of COVID-19, many researchers assume that each contact has the same probability of transmission (conditional on time and distance of contact), i.e., a binomial process (e.g., Jing et al. (2020); Kucharski et al. (2020)). Our results, however, suggest that this should be modelled as a two-step procedure when simulating contacts between individuals: First, it should be determined whether a case is infectious or not (i.e., a Bernoulli process), and second, conditional on being an infectious case, a binomial process should be used to represent actual transmission. This allows replicating the transmission dynamics of the virus more realistically and hence allows more realistic predictions from these models.

In conclusion, to the best of our knowledge, this study presents the results of the first nationwide register-based study on household transmission of COVID-19. These results are important as they show differences in transmission pattern with the age of both the primary case and the potential secondary cases within households. A large proportion of transmission was found to occur within households, highlighting the severity of COVID-19 transmission, and that preventative measures within households are urgently needed in order to prevent transmission. Moreover, monitoring the proportion of positive cases originating from households may be an important tool for public health authorities to measure community transmission.

## Data Availability

Data is available through Statens Serum Institut and The Danish Health Data Authority.

## Appendix A Overview of the COVID-19 Epidemic in Denmark

**Figure A.1:**
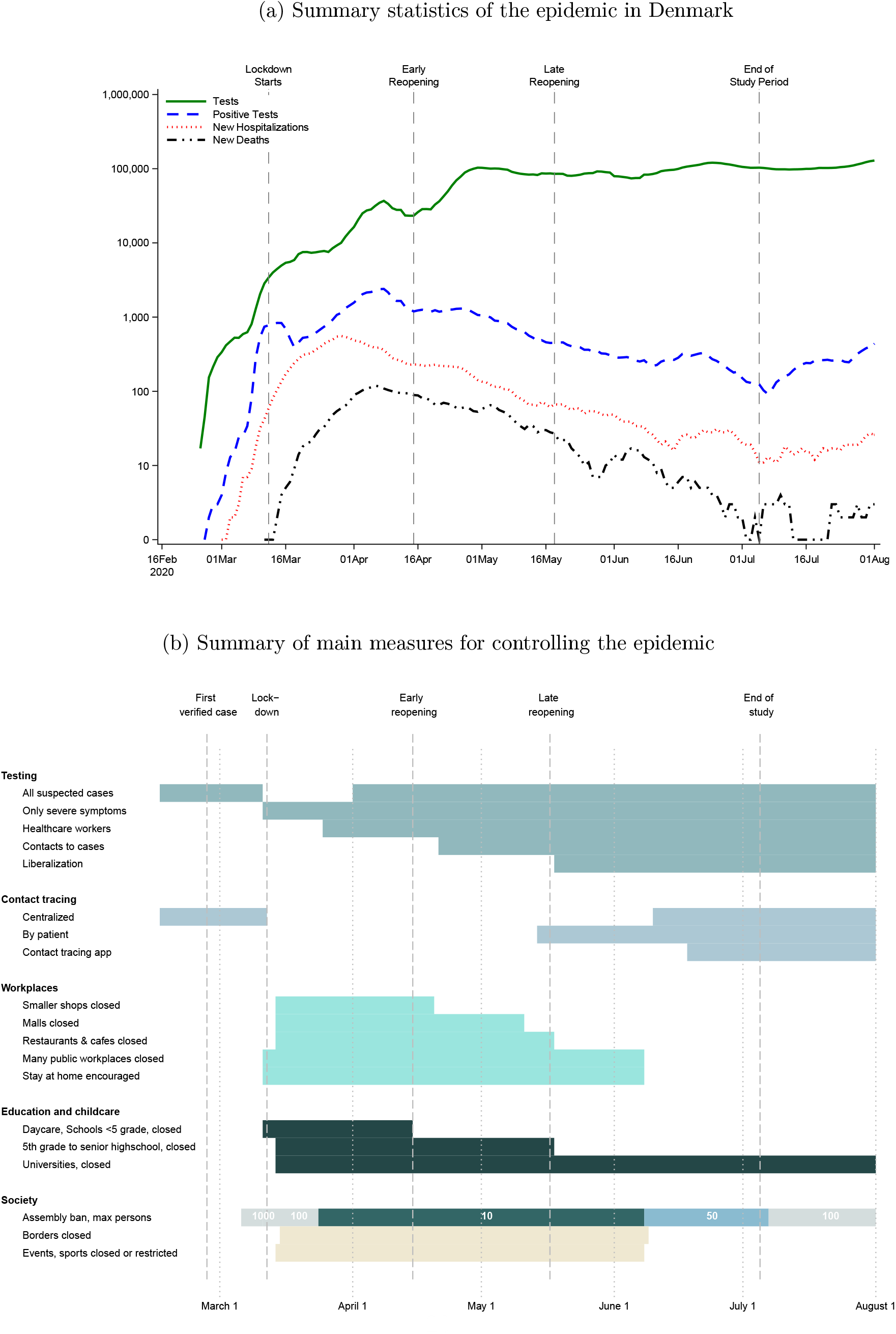
Overview of the Epidemic in Denmark

**Figure A.2:**
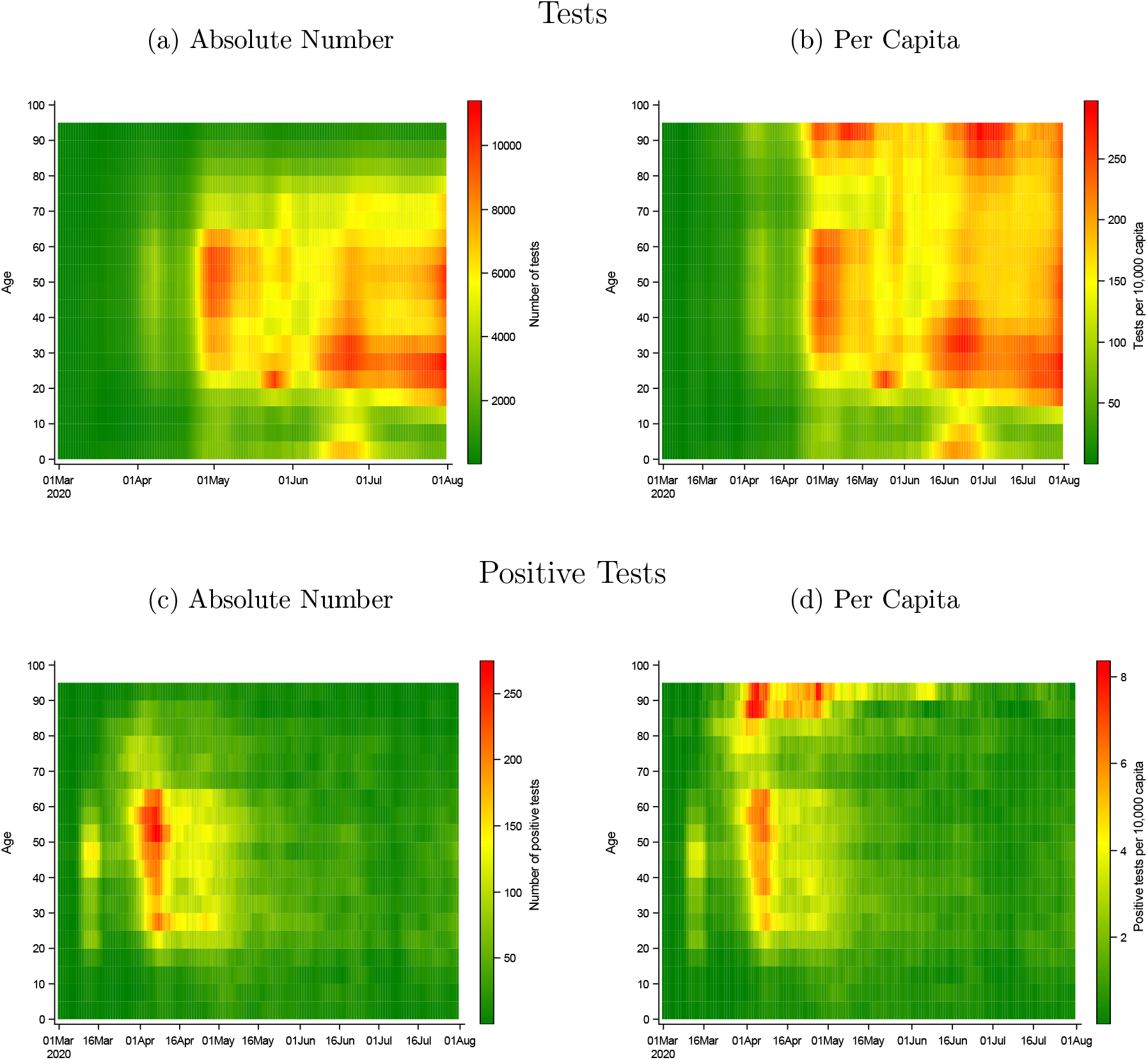
Age Specific Probability of Obtaining a Test and a Positive Test

Notes: Age is 5-year-age groups. Numbers represent a 7-day-rolling sum. The figure is inspired by Marc Bevand, https://github.com/mbevand/florida-covid19-line-list-data

**Table A.1:** Days From Test to Test Result

|  | Lockdown | Early Reopening | Late Reopening | Total |
| --- | --- | --- | --- | --- |
| 25 Percentile | 0 | 0 | 1 | 0 |
| 50 Percentile | 1 | 1 | 1 | 1 |
| 75 Percentile | 1 | 2 | 1 | 1 |
| 95 Percentile | 2 | 4 | 2 | 3 |
| Mean | 0.9 | 1.3 | 1.0 | 1.0 |
| Number of tests | 87,034 | 375,444 | 679,847 | 1,142,325 |

Notes: This table provides summary statistics on the number of days between the day of the test and the day of receiving the result (for all SARS-CoV-2 tests obtained in the study period).

## Appendix B Summary Statistics

**Table B.1:** Summary Statistics, Primary Cases

|  | Lockdown | Early Reopening | Late Reopening | Total |
| --- | --- | --- | --- | --- |
| **Total** | 4,727 | 2,907 | 1,267 | 8,901 |
| **Sex** |  |  |  |  |
| Male | 1,771 | 988 | 555 | 3,314 |
| Female | 2,950 | 1,918 | 712 | 5,580 |
| **Age** |  |  |  |  |
| 0 - 4 | 21 | 26 | 23 | 70 |
| 5 - 9 | 8 | 31 | 25 | 64 |
| 10 - 14 | 7 | 31 | 32 | 70 |
| 15 - 19 | 66 | 93 | 59 | 218 |
| 20 - 24 | 220 | 226 | 116 | 562 |
| 25 - 29 | 377 | 295 | 126 | 798 |
| 30 - 34 | 338 | 277 | 107 | 722 |
| 35 - 39 | 340 | 209 | 110 | 659 |
| 40 - 44 | 428 | 257 | 95 | 780 |
| 45 - 49 | 457 | 245 | 95 | 797 |
| 50 - 54 | 509 | 263 | 117 | 889 |
| 55 - 59 | 507 | 241 | 86 | 834 |
| 60 - 64 | 417 | 209 | 63 | 689 |
| 65 - 69 | 234 | 89 | 49 | 372 |
| 70 - 74 | 238 | 109 | 55 | 402 |
| 75 - 79 | 222 | 88 | 46 | 356 |
| 80 - 84 | 164 | 91 | 34 | 289 |
| 85 - 89 | 114 | 76 | 17 | 207 |
| 90 - 94 | 46 | 39 | 9 | 94 |
| 95 - 99 | 7 | 11 | <5 | 21 |
| 100 - 104 | <5 | - | - | <5 |
| **Household Members** |  |  |  |  |
| 1 | 1,115 | 727 | 277 | 2,119 |
| 2 | 1,650 | 917 | 368 | 2,935 |
| 3 | 728 | 477 | 216 | 1,421 |
| 4 | 766 | 473 | 240 | 1,479 |
| 5 | 358 | 234 | 124 | 716 |
| 6 | 110 | 79 | 42 | 231 |

Notes: This table provides summary statistics on the primary cases included in the study.

**Table B.2:** Summary Statistics, Potential Secondary Cases

|  | Lockdown | Early Reopening | Late Reopening | Total |
| --- | --- | --- | --- | --- |
| **Total** | 7,386 | 4,621 | 2,226 | 14,233 |
| **Sex** |  |  |  |  |
| Male | 4,016 | 2,683 | 1,123 | 7,822 |
| Female | 3,358 | 1,937 | 1,103 | 6,398 |
| **Age** |  |  |  |  |
| 0 - 4 | 614 | 476 | 221 | 1,311 |
| 5 - 9 | 736 | 454 | 230 | 1,420 |
| 10 - 14 | 874 | 490 | 225 | 1,589 |
| 15 - 19 | 914 | 477 | 183 | 1,574 |
| 20 - 24 | 503 | 372 | 174 | 1,049 |
| 25 - 29 | 449 | 324 | 157 | 930 |
| 30 - 34 | 312 | 293 | 143 | 748 |
| 35 - 39 | 304 | 259 | 143 | 706 |
| 40 - 44 | 393 | 264 | 155 | 812 |
| 45 - 49 | 439 | 271 | 150 | 860 |
| 50 - 54 | 493 | 284 | 153 | 930 |
| 55 - 59 | 446 | 252 | 92 | 790 |
| 60 - 64 | 345 | 176 | 68 | 589 |
| 65 - 69 | 210 | 86 | 42 | 338 |
| 70 - 74 | 154 | 71 | 48 | 273 |
| 75 - 79 | 107 | 36 | 27 | 170 |
| 80 - 84 | 53 | 28 | 14 | 95 |
| 85 - 89 | 26 | <5 | <5 | 31 |
| 90 - 94 | <5 | <5 | - | 5 |
| 95 - 99 | - | - | - | - |
| 100 - 104 | - | - | - | - |
| **Household Members** |  |  |  |  |
| 1 | - | - | - | - |
| 2 | 1,650 | 917 | 368 | 2,935 |
| 3 | 1,456 | 954 | 432 | 2,842 |
| 4 | 2,298 | 1,419 | 720 | 4,437 |
| 5 | 1,432 | 936 | 496 | 2,864 |
| 6 | 550 | 395 | 210 | 1,155 |

Notes: This table provides summary statistics on the potential secondary cases included in the study.

**Table B.3:**
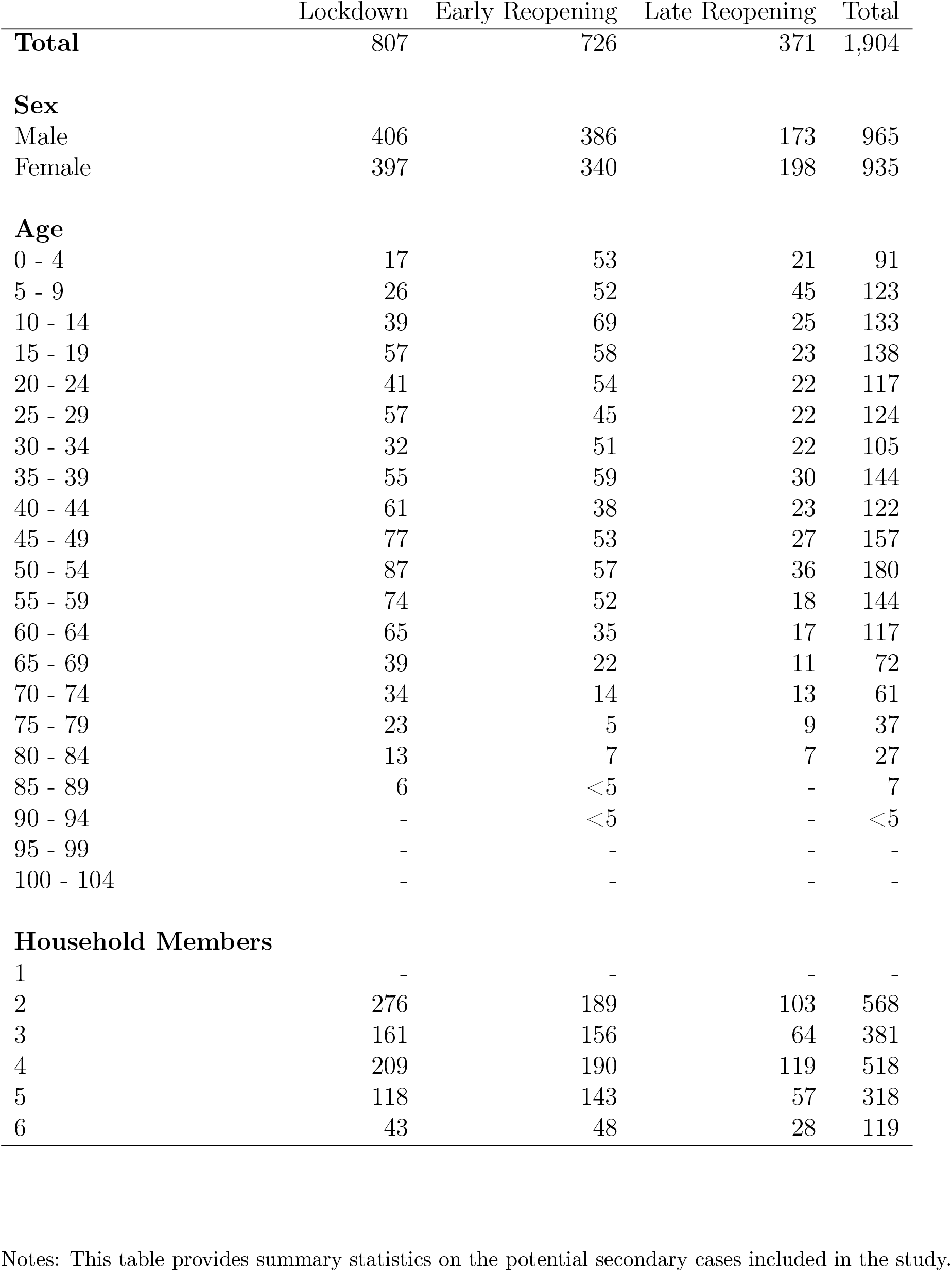
Summary Statistics, Positive Secondary Cases

**Figure B.1:**
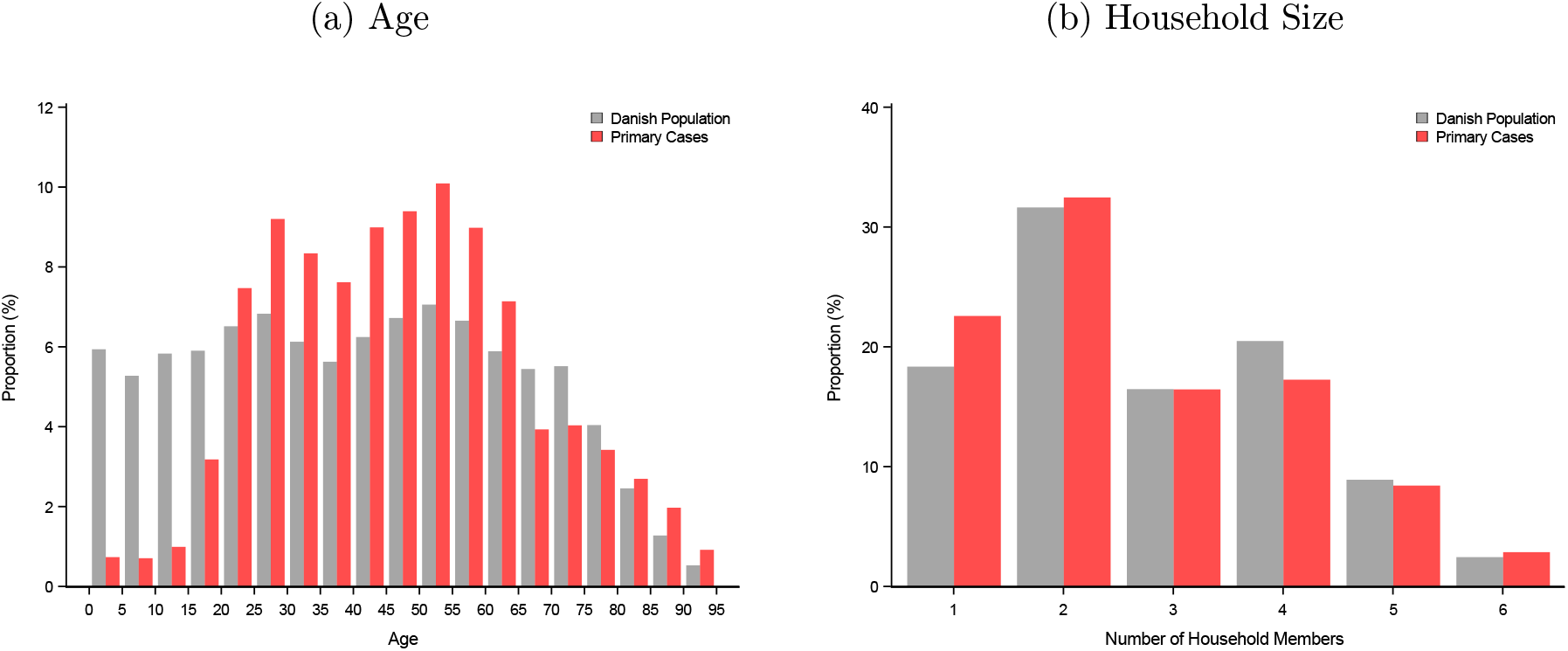
Descriptive Statistics of Primary Cases and Danish Population

Summary statistics of the primary cases and the full Danish population by age, sex, and household size, from households with maximal six persons. (a) shows the distribution of all individuals in Denmark (gray) and the distribution of primary cases (red) by 5-year age groups. (b) shows the household size for primary cases (red) and the overall population (gray).

## Appendix C Age Structured Attack Rate

**Figure C.1:**
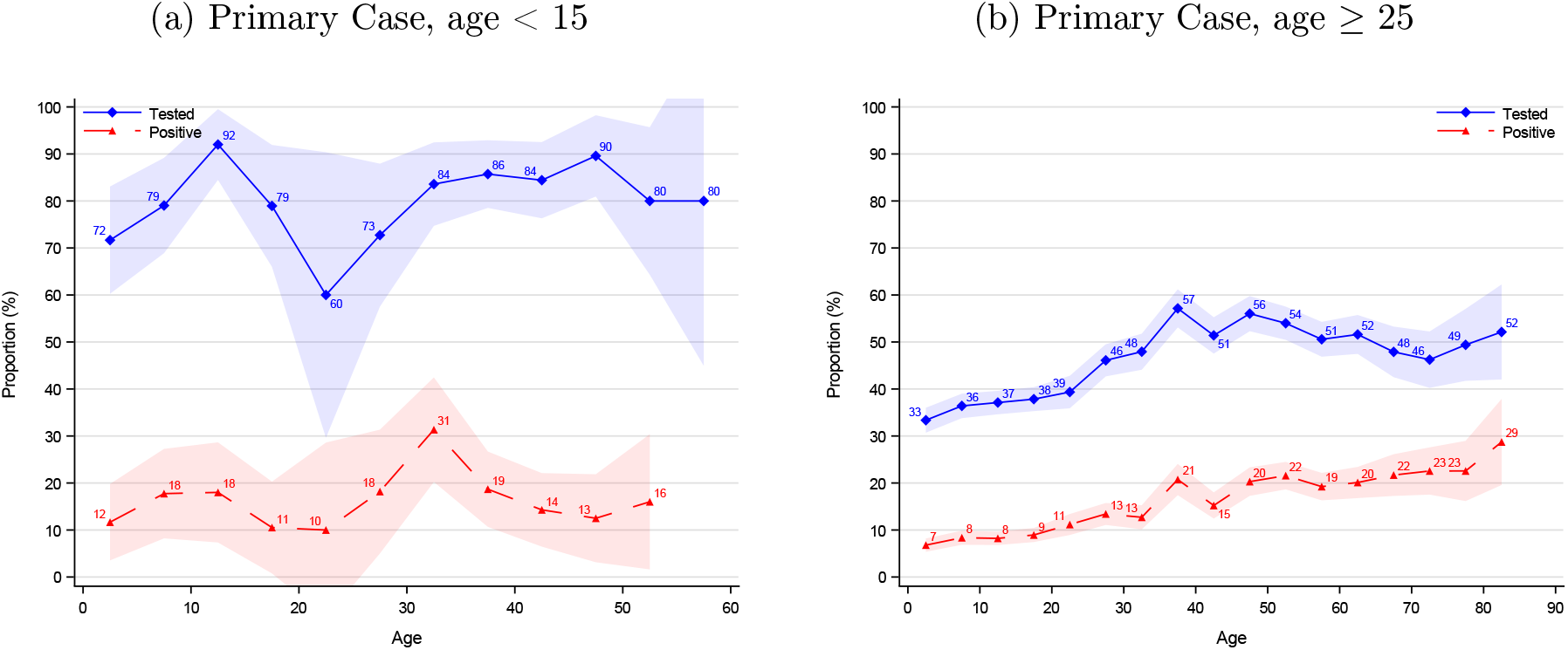
Age Structured Attack Rate by Age of Primary Case

Notes: Shaded areas are 95% confidence bands clustered on the household level.

**Figure C.2:**
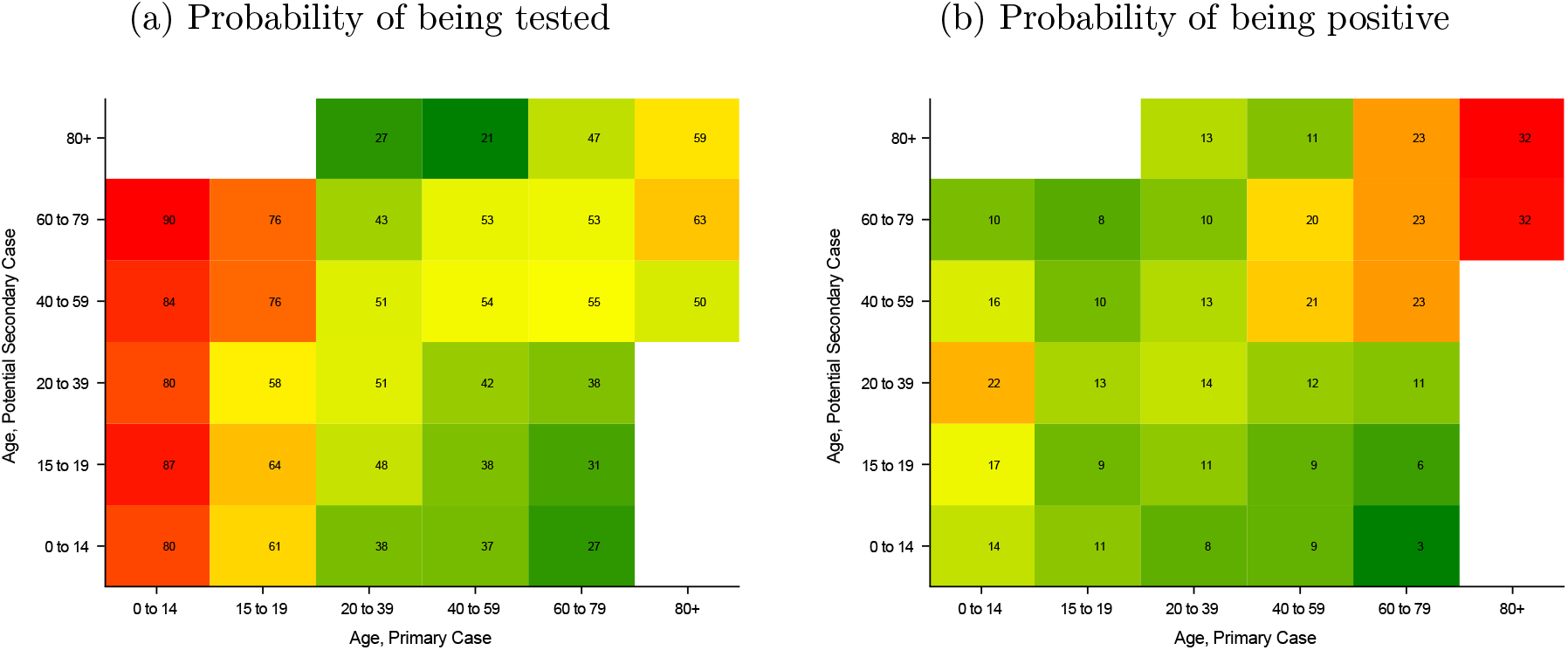
Age Structured Attack Rate and Transmission Risk(a) Probability of being tested(b) Probability of being positive

Notes: Panel a shows the probability of being tested and panel b the probability of having a positive test. The age of the primary case is depicted on the x-axis and the age of potential secondary cases on the y-axis.

### Appendix C.1 Age structured Attack Rate by Sex

**Figure C.3:**
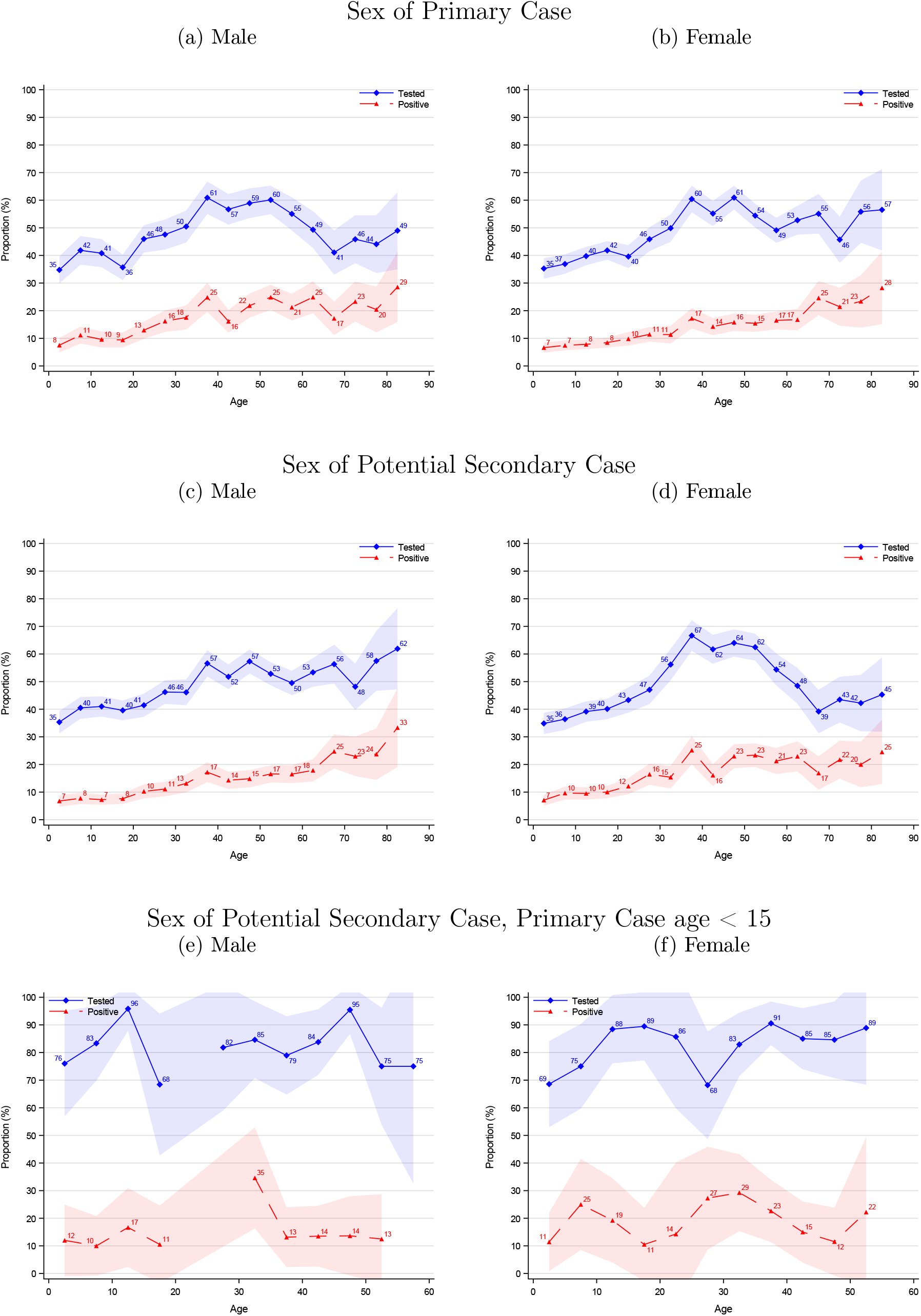
Age Structured Attack Rate by Sex

## Appendix D Dynamics over the epidemic

### Appendix D.1 Testing Dynamics over the epidemic

Figure D.1 panel a-c presents the testing dynamics for the three periods of the epidemic. Panel a shows that during lockdown only few percent of the other household members were tested and that there was a high positive rate of the ones tested. Panel b shows that during the early reopening, more household members were being tested and the positive rate declined accordingly. Lastly, panel c shows that in the late reopening even more household members were being tested and the positive rate declined further. Panel d-f shows the proportion of household members ever being tested and ever being positive. During lockdown 25% of secondary household members were tested after 14 days, during early reopening 65% were tested, and during late reopening 82% were tested. Similarly, 11% had tested positive after 14 days during lockdown, 16% during early reopening and 17% during late reopening. Another aspect of testing is how long a person has to wait for the result after obtaining the test. Table A.1 shows that 75% of all cases received the result within 1 day during lockdown, within 2 days during early reopening, and within 1 day during late reopening.

**Figure D.1:**
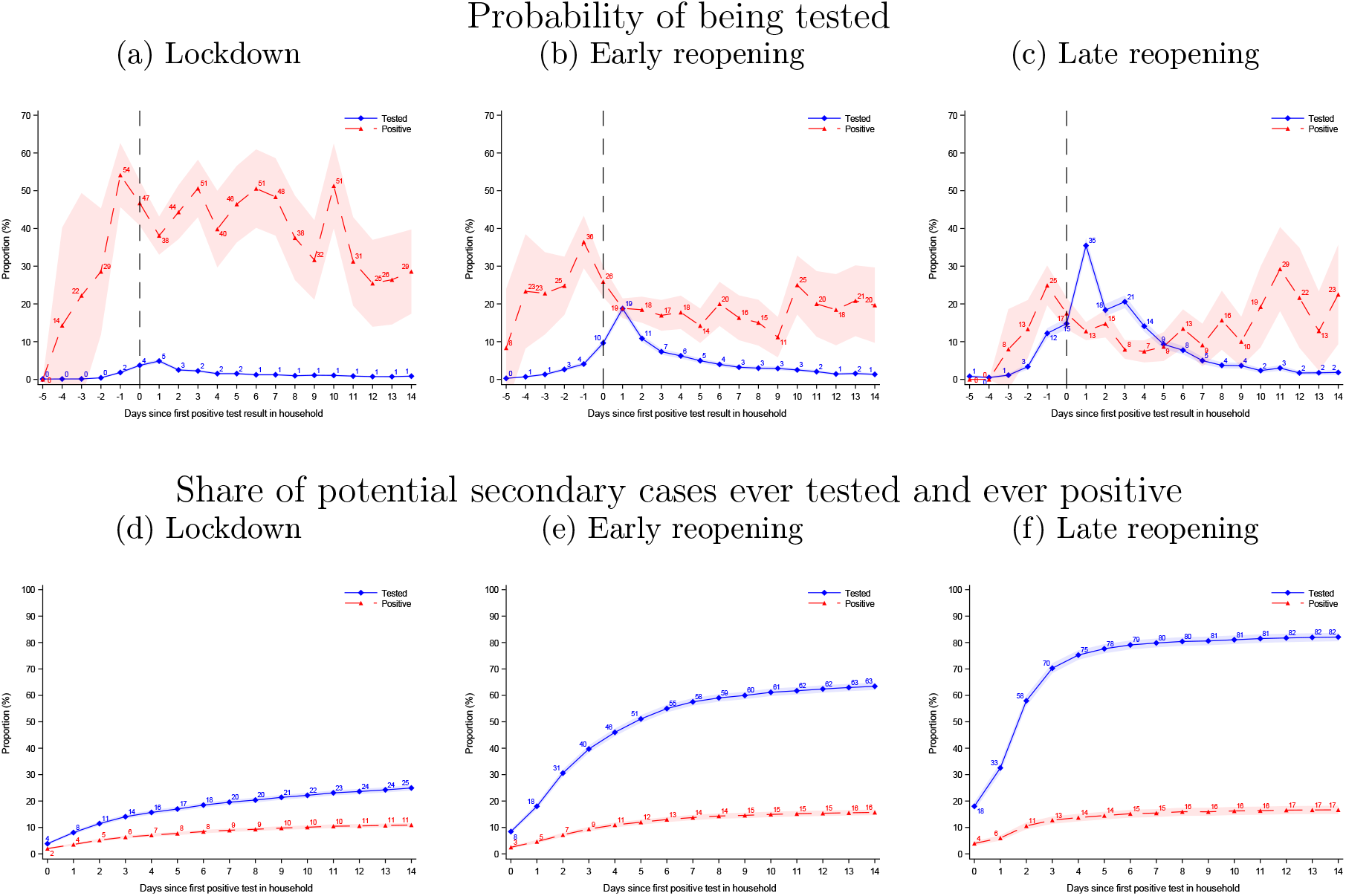
Testing Dynamics over the epidemic

Notes: Shaded areas are 95% confidence bands clustered on the household level.

### Appendix D.2 Age Structured Attack Rate over the epidemic

Figure D.2 illustrates the age structured attack rate separately for the three periods of the epidemic. The testing rate clearly increases during the epidemic, while the rate of positive test results is relatively constant. Both rates increase with age of the case.

**Figure D.2:**
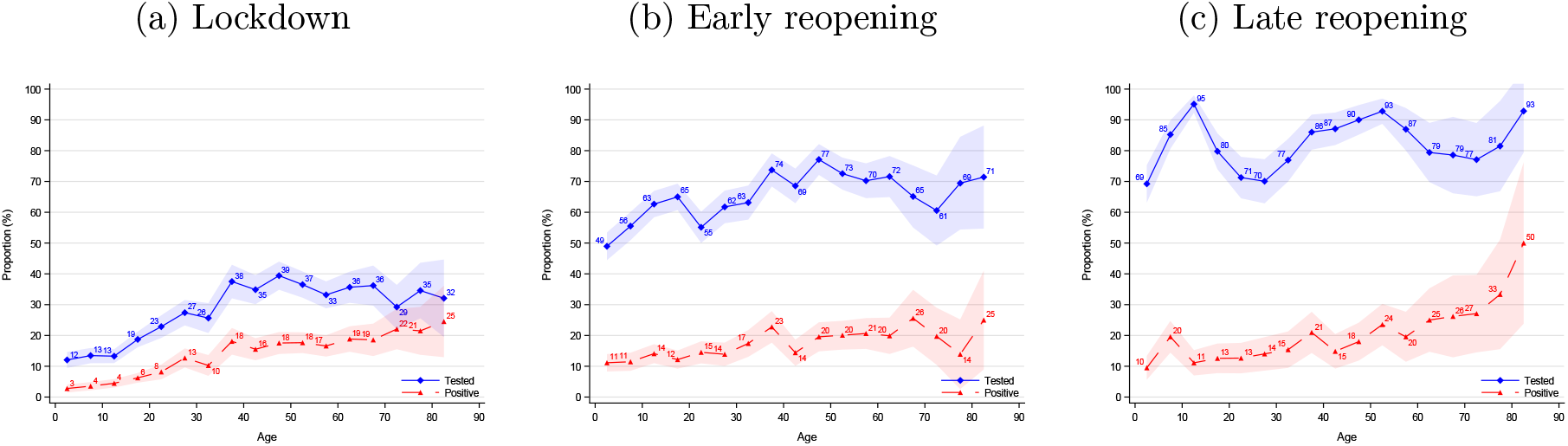
Age Structured Attack Rate over the epidemic(a) Lockdown(b) Early reopening(c) Late reopening

Notes: This figure illustrates the age structured attack rate separately for the three periods of the epidemic. Figure 3 shows the estimates for the pooled analysis. The testing rate clearly increases during the epidemic, while the positive rate is fairly constant. Shaded areas are 95% confidence bands clustered on the household level.

### Appendix D.3 Household Attack Rate over the epidemic

Figure D.3 presents the results for the number of individuals infected across each household size for the three periods of the epidemic. The results are relatively constant over the three periods.

**Figure D.3:**
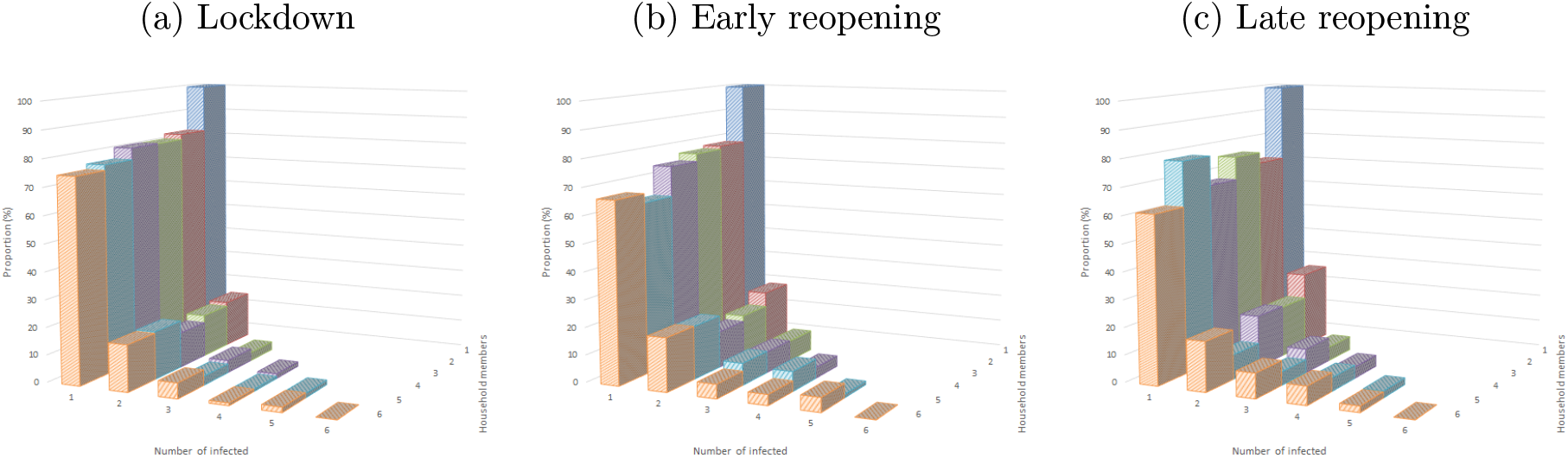
Attack Rate within the Household

Notes: Figure 4 illustrates the pooled estimate for the three periods.

## Appendix E Regression Estimates

**Table E.1:** Regression Estimates: Attack Rate

|  | I | II | III | IV |
| --- | --- | --- | --- | --- |
| Intercept | 0.1336<br>(0.0035)** | 0.1667<br>(0.0099)** | 0.1556<br>(0.0102)** | 0.1556<br>(0.0123)** |
| Lockdown |  | -0.0578<br>(0.0107)** | -0.0569<br>(0.0107)** | -0.0538<br>0.0133** |
| Early Reopening |  | -0.0095<br>(0.0119) | -0.0078<br>(0.0119) | -0.0123<br>(0.0144) |
| Late Reopening |  | -<br>(-) | -<br>(-) | -<br>(-) |
| Female |  |  | 0.0224<br>(0.0057)** | 0.0225<br>(0.0161) |
| Female × Lockdown | | | | -0.0068<br>(0.0177) |
| Female × Early Reopening | | | | 0.0107<br>(0.0194) |
| Number of Households | 6,782 | 6,782 | 6,782 | 6,782 |
| Number of Observations | 14,220 | 14,220 | 14,220 | 14,220 |

Notes: Standard errors in parenthesis, clustered on the household level. \*p<0.05, \*\*p<0.01.

**Table E.2:** Regression Estimates: Attack Rate dependent on age

|  | I | II | III |
| --- | --- | --- | --- |
| Intercept | 0.0939<br>(0.0107)** | 0.1012<br>(0.0162)** | 0.0858<br>(0.0182)** |
| Age | 0.0024<br>(0.0001)** | 0.0021<br>(0.0004)** | 0.0022<br>(0.0004)** |
| Lockdown | 0.0600<br>(0.0107)** | -0.0820<br>(0.0172)** | -0.0771<br>(0.0193)** |
| Early Reopening | -0.0079<br>(0.0119) | 0.0066<br>(0.0193) | 0.0032<br>(0.0213) |
| Late Reopening | (-)<br>(-) | (-)<br>(-) | (-)<br>(-) |
| Age × Lockdown | | 0.0007<br>(0.0005) | 0.0007<br>(0.0005) |
| Age × Early Reopening | | -0.0005<br>(0.0005) | -0.0005<br>(0.0005) |
| Female |  |  | 0.0282<br>(0.0161) |
| Female × Lockdown | | | -0.0069<br>(0.0175) |
| Female × Early Reopening | | | 0.0109<br>(0.0109) |
| Number of Households | 6,782 | 6,782 | 6,782 |
| Number of Observations | 14,220 | 14,220 | 14,220 |

Notes: Specification (III) shows that individuals have a baseline probability of getting infected of 8.58%. The probability increases by 0.22 pp for each patient year of age. Thus a 10-year-old has a 10.2% risk, a 30-year-old has 15.0%, and a 60-year-old has 22.2%, according to the model. Standard errors in parenthesis, clustered on the household level. \* $p < 0.05$ , \*\* $p < 0.01$ .

**Table E.3:** Proportion of Cases by Household Size

|  |  | Number of Household Members |  |  |  |  |  |
| --- | --- | --- | --- | --- | --- | --- | --- |
|  |  | 1 | 2 | 3 | 4 | 5 | 6 |
| Number of cases | 1 | 100.0<br>(.) | 80.6<br>(0.73) | 78.6<br>(1.09) | 76.7<br>(1.10) | 72.8<br>(1.66) | 69.7<br>(3.02) |
|  | 2 |  | 19.4<br>(0.73) | 16.0<br>(0.97) | 14.5<br>(0.91) | 16.6<br>(1.39) | 18.2<br>(2.54) |
|  | 3 |  |  | 5.4<br>(0.60) | 6.0<br>(0.62) | 5.6<br>(0.86) | 6.1<br>(1.57) |
|  | 4 |  |  |  | 2.8<br>(0.43) | 3.5<br>(0.69) | 3.0<br>(1.13) |
|  | 5 |  |  |  |  | 1.5<br>(0.46) | 3.0<br>(1.13) |
|  | 6 |  |  |  |  |  | 0.0<br>(.) |
| Number of observations |  | 2,119 | 5,870 | 4,263 | 5,916 | 3,580 | 1,386 |
| Number of households |  | 2,119 | 2,935 | 1,421 | 1,479 | 716 | 231 |

Notes: Estimates are in percentages. Standard errors in parenthesis, clustered on the household level.

**Table E.4:** Proportion of Secondary Cases Per Infected Household

|  | Number of Secondary of Cases |  |  |  |  |  |
| --- | --- | --- | --- | --- | --- | --- |
|  | 0 | 1 | 2 | 3 | 4 | 5 |
|  | 77.0<br>(0.56) | 16.7<br>(0.48) | 5.8<br>(0.39) | 3.1<br>(0.36) | 2.0<br>(0.46) | 0.0<br>(.) |
| Number of observations | 21,015 | 21,015 | 15,145 | 10,882 | 4,966 | 1,386 |
| Number of households | 6,782 | 6,782 | 3,847 | 2,426 | 947 | 231 |

Notes: The estimates are calculated from households with at least one potential secondary case, i.e., households with two to six members. The estimate for two secondary cases is calculated from households with at least two potential secondary cases, i.e., households with three to six members. Estimates are in percentages. Standard errors in parenthesis, clustered on the household level.

## Appendix F Robustness for Definition of Co-Primary Cases

**Table F.1:** Robustness for Definition of Co-Primary Cases: Attack Rate

|  | I | II | III | IV | V |
| --- | --- | --- | --- | --- | --- |
| Excluding days ≤ | None | 0 | 1 | 2 | 3 |
| Intercept | 0.1667<br>(0.0099)** | 0.1328<br>(0.0091)** | 0.1133<br>(0.0087)** | 0.0692<br>(0.0070)** | 0.0453<br>(0.0056)** |
| Lockdown | -0.0578<br>(0.0107)** | -0.0419<br>(0.0099)** | -0.0371<br>(0.0094)** | -0.0092<br>(0.0077) | 0.0034<br>(0.0064) |
| Early Reopening | -0.0095<br>(0.0119) | 0.0027<br>(0.0111) | 0.0027<br>(0.0105) | 0.0224<br>(0.0088)* | 0.0245<br>(0.0074)** |
| Late Reopening | -<br>(-) | -<br>(-) | -<br>(-) | -<br>(-) | -<br>(-) |
| Number of Households | 6,782 | 6,636 | 6,545 | 6,413 | 6,336 |
| Number of Observations | 14,220 | 13,871 | 13,610 | 13,271 | 13,036 |

Notes: This table provides estimates on the robustness of the estimates for the attack rate depending on the definition of co-primary cases. Column (I) corresponds to column (II) in Table E.1. In column (II) we exclude secondary cases that test positive on the same day ( $t = 0$ ) as the primary case. In column (III) we exclude secondary cases that test positive within one day ( $t \leq 1$ ) of the primary case. In column (V) we exclude secondary cases that test positive within three days ( $t \leq 5$ ) of the primary case. Standard errors in parenthesis, clustered on the household level. \* $p < 0.05$ , \*\* $p < 0.01$ .

**Figure F.1:**
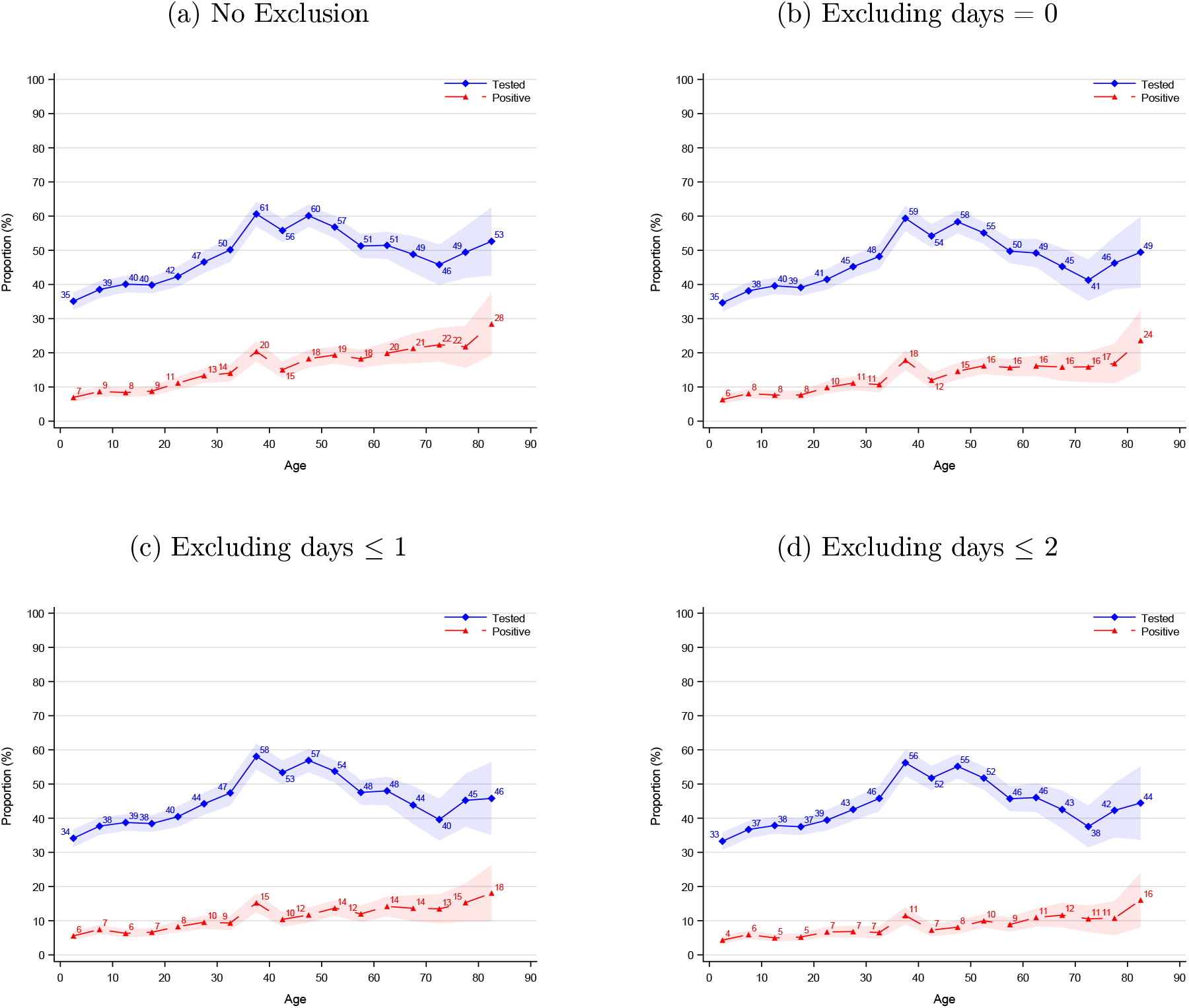
Robustness for Definition of Co-Primary Cases: Age Structured Attack Rate

Notes: This figure illustrates robustness for the definition of co-primary cases with respect to the age structured attack rate. Panel a has no restrictions and is the same as figure 3. Panel b excludes secondary cases testing positive the same day as the primary case (*t* = 0). Panel c excludes secondary cases testing positive within 1 day of the primary case (*t* ≤1). Panel d excludes secondary cases testing positive within 2 days of the primary case (*t* ≤2).

## Appendix G Members of the Expert Group for Mathematical Modelling of COVID-19 at Statens Serum Institut, Denmark

- Robert Leo Skov, Statens Serum Institut
- Kaare Græsbøll, Department of Applied Mathematics and Computer Science, Technical University of Denmark
- Lasse Engbo Christiansen, Department of Applied Mathematics and Computer Science, Technical University of Denmark
- Sune Lehmann, Department of Applied Mathematics and Computer Science, Technical University of Denmark
- Uffe Høgsbro Thygesen, Department of Applied Mathematics and Computer Science, Technical University of Denmark
- Jonas Lybker Juul, Department of Applied Mathematics and Computer Science, Technical University of Denmark
- Carsten Thure Kirkeby, Department of Veterinary and Animal Sciences, Faculty of Health and Medical Sciences, University of Copenhagen
- Matt Denwood, Department of Veterinary and Animal Sciences, Faculty of Health and Medical Sciences, University of Copenhagen
- Tariq Halasa, Department of Veterinary and Animal Sciences, Faculty of Health and Medical Sciences, University of Copenhagen
- Theis Lange, Section of Biostatistics, University of Copenhagen
- Kim Sneppen, Niels Bohr Institute, University of Copenhagen
- Lone Simonsen, Roskilde University
- Viggo Andreasen, Roskilde University
- Anders Perner, Rigshospitalet
- Laust Hvas Mortensen, Statistics Denmark
- Søren Mikael Neermark, Danish Health Authority
- Mathias Heltberg, Niels Bohr Institute, University of Copenhagen, & Statens Serum Institut
- Frederik Plesner Lyngse, Department of Economics & Center for Economic Behaviour and Inequality, University of Copenhagen, & Danish Ministry of Health & Statens Serum Institut
- Peter Michael Bager, Statens Serum Institut
- Tyge Arnold Larsen, Statens Serum Institut & Danish Ministry of Health
- Pernille Skorstengaard, Statens Serum Institut & Danish Ministry of Health
- Christine Foltmar Gammelgaard, Statens Serum Institut & Danish Ministry of Health
- Esther Løffler, Statens Serum Institut & Danish Ministry of Health

## Footnotes

* We thank Statens Serum Institut and The Danish Health Data Authority for data access and helpful institutional knowledge. We also thank the rest of the Expert Group for Mathematical Modelling of COVID-19 at Statens Serum Institut (Appendix G) for helpful discussion and comments.

## Notes

### Competing Interest Statement

The authors have declared no competing interest.

### Funding Statement

Frederik Plesner Lyngse: Independent Research Fund Denmark (Grant no. 9061-00035B.); Novo Nordisk Foundation (grant no. NNF17OC0026542); the Danish National Research Foundation through its grant (DNRF-134) to the Center for Economic Behavior and Inequality (CEBI) at the University of Copenhagen.
Carsten Thure Kirkeby: None.
Tariq Halasa: None.
Viggo Andreasen: Carlsberg Foundation, Semper Ardens Research Project, CF20-0046.
Robert Leo Skov: None.
Tyra Grove Krause: None.
Frederik Trier Møller: None.
Kåre Mølbak: None.

### Author Declarations

Statens Serum Institut and The Danish Health Data Authority.

